# Rate of tuberculosis screening and isoniazid prophylaxis treatment among human immunodeficiency virus patients in Obio/Akpor Local Government Area: A retrospective study

**DOI:** 10.1101/2023.01.26.23285062

**Authors:** Precious Ruth Edoja, Uchechukwu Ifeanyichukwu Apugo, Ekenedilichukwu Chukwudubem Anekwe

## Abstract

**Background:** The co-occurrence of Tuberculosis (TB) and Human Immunodeficiency Virus (HIV) has been recognised as a global public health problem with considerable mutual interaction and a leading cause of death worldwide.

**Aim:** To determine the rate of TB screening among People Living with HIV (PLWH) and treatment with isoniazid (INH) prophylaxis in directly observed treatment short course (DOTS) clinics in Primary Healthcare centres (PHCs) in Obio/Akpor Local Government Area of Rivers state.

**Materials and Method:** This clinic-based, two-year retrospective cross-sectional study involved a complete review and abstraction of all records for HIV patients (18 years and above) who have been receiving HIV care and treatment for at least 6 months prior to the study in DOTS clinics in five selected PHCs in OBALGA. The data were extracted using a checklist while the statistical analysis of the study was carried out using SPSS version 23.

**Results:** The proportion of HIV-positive patients screened for TB was 79.8% while those who had TB diagnostic evaluation among them was 38.6%, and 38.7% of the HIV/TB co-infected patients were placed on INH prophylaxis.

**Conclusion:** Insufficient attention is being paid to TB diagnostic evaluation and IPT. Hence, it is recommended that the PHCs in the LGA should be equipped with appropriate devices for TB diagnosis as well as engage the Healthcare Workers in sensitization workshops on the need for continuous screening of PLWH for TB.

## INTRODUCTION

Tuberculosis (TB) and Human Immunodeficiency Virus / Acquired immune deficiency syndrome (HIV/AIDS) are the two leading causes of death from infectious diseases worldwide (Pathmanathan et al., 2017), while their co-occurrence is recognised as a global public health problem with considerable mutual interaction and a leading cause of death worldwide (de Colombani et al., 2004; Igbokwe, Abugu & Aji, 2020). They both have a strong fatal bidirectional correlation or interaction where each drives the progress or potentiates the adverse effects of the other (Makanjuola, Taddese & Boot, 2014). Hence, their co-existence can be regarded as a syndemic (the convergence of two or more diseases that act synergistically to magnify the burden of disease) interaction (Kwan & Ernst, 2011). Also, co-infection affects the prognosis of patients and complicates clinical diagnosis and management plans through an unusual presentation of symptoms, adverse drug reactions, drug–drug interactions, and overlapping drug toxicities between anti-TB drugs and highly active antiretroviral therapy (ART) (Pawlowski et al., 2012; Ayele et al., 2017).

Alone, TB is identified as the most prevalent disease and the major cause of death with a specific aetiology among People Living with HIV (PLWH), including those on ART (Getahun et al., 2010; Igbokwe, Abugu & Aji, 2020), as it accounts for about 25% of all causes of death among HIV patients (Getahun et al., 2010; WHO, 2017). On the other hand, HIV is seen as the most potent force driving the TB epidemic in countries with a high prevalence of HIV (de Colombani et al., 2004). TB/HIV coinfection presents as one of the major challenges of global TB control with reports showing that TB is the most common life-threatening and leading cause of death, while the World Health Organisation (WHO) estimated that among the 8.6 million people who developed TB, about 1.1 million new TB cases (13% of total TB cases) were reported among PLWH (WHO, 2012; Karo et al., 2014).

Given the high rate of TB in HIV-infected individuals, as well as their close interactions, the WHO developed a policy on collaborative TB/HIV activities as part of the core HIV and TB prevention, care, and treatment services (WHO, 2004). These include interventions aimed at reducing TB burdens among PLWH, such as the scale-up of ART, Active Case Finding (ACF), and initiation of Isoniaziad preventive therapy (IPT) among TB/HIV co-infected patients (WHO, 2008b; Igbokwe, Abugu & Aji, 2020). According to Giri et al (2013), all HIV-infected individuals should be tested for TB prior to ART initiation in countries where TB is endemic. Though these interventions are deemed very essential as they will lead to the reduction in the risk of TB progression and transmission among both PLWH and those without HIV infection (Kanabus, 2017), co-infection of TB/HIV has continued to be a leading cause of morbidity and mortality among PLWH as millions of patients go unreported every year, especially in developing countries and Sub-Saharan African (SSA) countries like Nigeria (WHO, 2015b). WHO also estimated that Nigeria has a high burden of both TB and HIV (WHO, 2015a), as well as ranked 4th in TB infection worldwide, with over 80% of TB cases in the country still undetected (WHO, 2016). Another report by the Nigeria Tuberculosis Profile in 2017 revealed that Nigeria is placed third in the incidence of TB/HIV co-infection globally, with poor case finding due to reliance on passive case finding, missed opportunities to scale-up of TB screening among PLHW and provision of adequate care and treatment to PLWH diagnosed with TB implicated in this occurrence (WHO, 2015a). Hence, this study was aimed at determining the rate of screening for TB among PLWH, and treatment with isoniazid (INH) prophylaxis in directly observed treatment short course (DOTS) clinics in Primary Healthcare centres (PHCs) in Obio/Akpor LGA (OBALGA), Rivers state.

## METHODOLOGY

### Study Design and setting

This clinic-based study employed a retrospective cross-sectional design approach and involved a complete review and abstraction of all records for HIV cases in DOTS clinics in five selected PHCs in OBALGA.

### Study Population

The study population was drawn from data of adult HIV-positive patients (18 years and above) who have been receiving HIV care and treatment for at least 6 months prior to the study and enrolled in five randomly selected DOTS clinics in OBALGA during a two-year period between January 2019 through December 2020.

### Study Instruments

All medical records and clinical data from adult HIV patients receiving care at the selected DOTS centres in the LGA were collected using a checklist adapted from the WHO Questionnaire/standardized forms prepared for monitoring and evaluation of TB/HIV activities (WHO, 2011). This was also used in the studies of Pasipamire et al. (2016), Geleto, Abate and Egata (2017) and Beshaw et al (2021). The checklist collected information on the basic socio-demographic data of the registered patients, their laboratory and medication history/ information as well as information on the indicators of ICF and IPT (TB screen done, TB screen result, diagnostic evaluation done, result of diagnostic evaluation, TB treatment started and ever received a course of IPT).

### Statistical Analysis

Data obtained were manually sorted and fed into the computer in an Excel spreadsheet where they were checked for completeness before analysis using Statistical Package for the Social Science (SPSS) version 23. Descriptive statistics were analysed and presented in charts and tables as frequency count and percentage.

### Ethical Consideration

Ethical approval was obtained from the Ethical Review Committee of University of Port Harcourt (reference number: UPH/CEREMAD/REC/MM74/064), while official permission to review the TB register was obtained from the Rivers State Ministry of Health and the Rivers State Primary Healthcare Management Board.

## RESULTS

### Socio-demographic and medical details of PLWH

A total of 925 patient folders were reviewed. According to the result as presented in table 1 below, the majority of the patients (40.1%) were aged 31 – 30, females (67.9%), married (63.1%), educated to secondary level (62.7%), self-employed (98.9%), Christians (98.9%) and lived in urban areas (90.5%). Analysis of the medical history of the patients revealed that the majority (95.0%) were categorised to be in the WHO clinical stage I, had CD4 cell count less than 350 (74.4%), already on ART (99.8%), for 2 – 5 years (73.1%) and on IPT regimen for 5 – 6 months (75.1%)

**Table 1:**
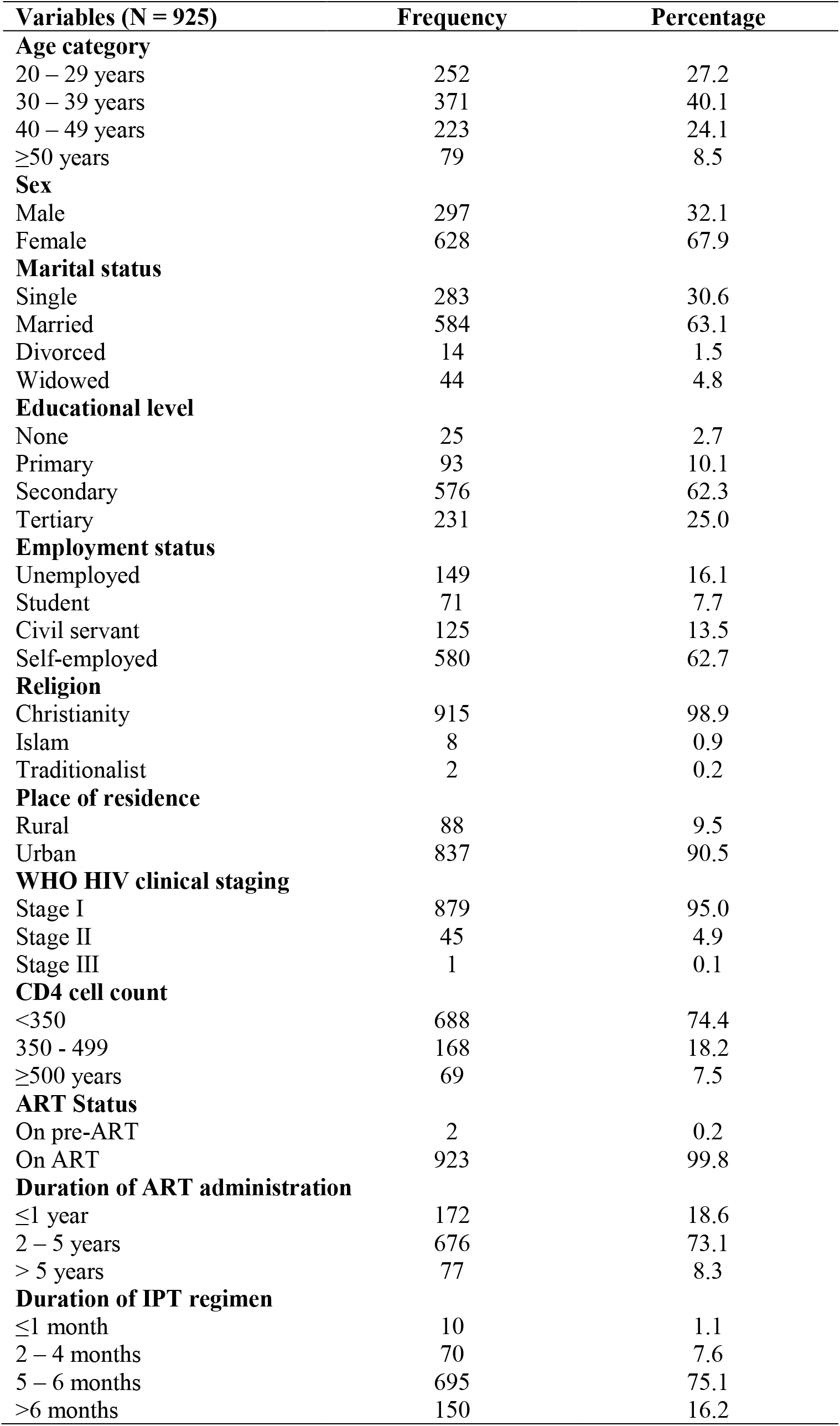
Socio-demographic and medical details of the PLWH in Obio/Akpor

| **Variables (N = 925)** | **Frequency** | **Percentage** |
| --- | --- | --- |
| **Age category** |  |  |
| 20 – 29 years | 252 | 27.2 |
| 30 – 39 years | 371 | 40.1 |
| 40 – 49 years | 223 | 24.1 |
| ≥50 years | 79 | 8.5 |
| **Sex** |  |  |
| Male | 297 | 32.1 |
| Female | 628 | 67.9 |
| **Marital status** |  |  |
| Single | 283 | 30.6 |
| Married | 584 | 63.1 |
| Divorced | 14 | 1.5 |
| Widowed | 44 | 4.8 |
| **Educational level** |  |  |
| None | 25 | 2.7 |
| Primary | 93 | 10.1 |
| Secondary | 576 | 62.3 |
| Tertiary | 231 | 25.0 |
| **Employment status** |  |  |
| Unemployed | 149 | 16.1 |
| Student | 71 | 7.7 |
| Civil servant | 125 | 13.5 |
| Self-employed | 580 | 62.7 |
| **Religion** |  |  |
| Christianity | 915 | 98.9 |
| Islam | 8 | 0.9 |
| Traditionalist | 2 | 0.2 |
| **Place of residence** |  |  |
| Rural | 88 | 9.5 |
| Urban | 837 | 90.5 |
| **WHO HIV clinical staging** |  |  |
| Stage I | 879 | 95.0 |
| Stage II | 45 | 4.9 |
| Stage III | 1 | 0.1 |
| **CD4 cell count** |  |  |
| <350 | 688 | 74.4 |
| 350 - 499 | 168 | 18.2 |
| ≥500 years | 69 | 7.5 |
| **ART Status** |  |  |
| On pre-ART | 2 | 0.2 |
| On ART | 923 | 99.8 |
| **Duration of ART administration** |  |  |
| ≤1 year | 172 | 18.6 |
| 2 – 5 years | 676 | 73.1 |
| > 5 years | 77 | 8.3 |
| **Duration of IPT regimen** |  |  |
| ≤1 month | 10 | 1.1 |
| 2 – 4 months | 70 | 7.6 |
| 5 – 6 months | 695 | 75.1 |
| >6 months | 150 | 16.2 |

### The proportion of HIV-positive patients Screened for TB

The study also analysed the proportion of patients who tested positive for HIV that were screened for TB, with the result presented in figure 1 below. According to the chart, more of the PLWH, 738 (79.8%) were screened for TB while 187 (20.2%) were not screened. Out of the 187 who were not screened, the data showed that 35 (3.8%) already tested positive for TB before undergoing the HIV test. Hence, only 152 (16.4%) persons who tested positive for HIV were not screened for TB.

**Figure 1:**
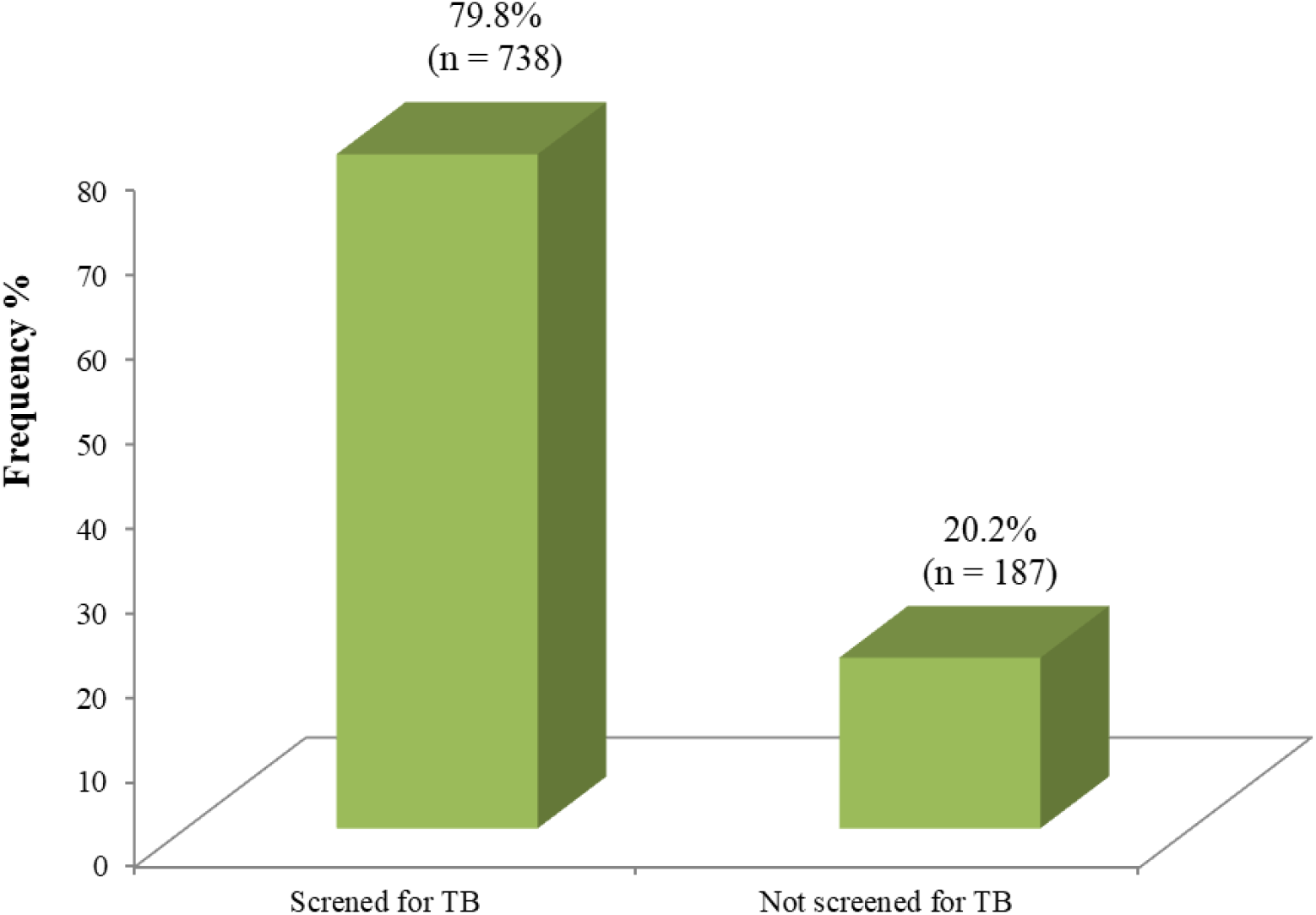
Distribution of PLWHA in Obio-Akpor screened for TB

Furthermore, the proportion of the HIV-positive patients who were screened for TB that had TB diagnostic evaluation was also determined and presented in figure 2 below. According to the result presented in the chart, out of the 738 patients screened for TB, 527 (71.4%) were not exposed to TB diagnostic evaluation while only 211 (38.6%) had a diagnostic evaluation done. Also, among the patients that had the diagnostic evaluation done, 60 (28.4%) were discovered to be positive for TB while 151 (71.6%) had negative TB evaluation.

**Figure 2:**
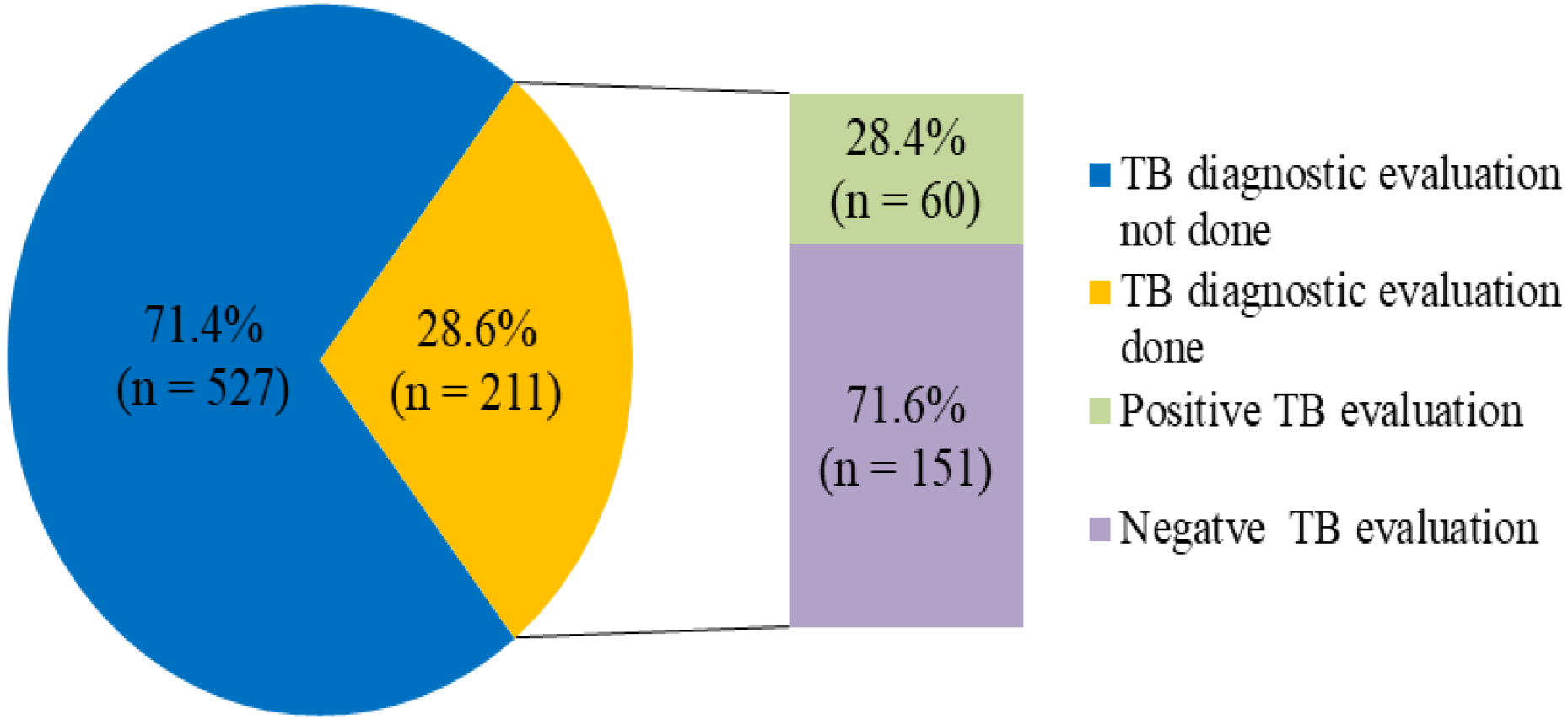
TB diagnostic evaluation among PLWH in Obio/Akpor

### The proportion of HIV/TB Positive patients placed on INH prophylaxis

The study also analysed the proportion of the PLWH who had positive TB evaluation that was placed on INH prophylaxis. According to the result presented in figure 3 below, out of the 60 PLWH that tested positive for TB evaluation, 22 (38.7%) were placed on INH prophylaxis while 38 (63.3%) were not.

**Figure 3:**
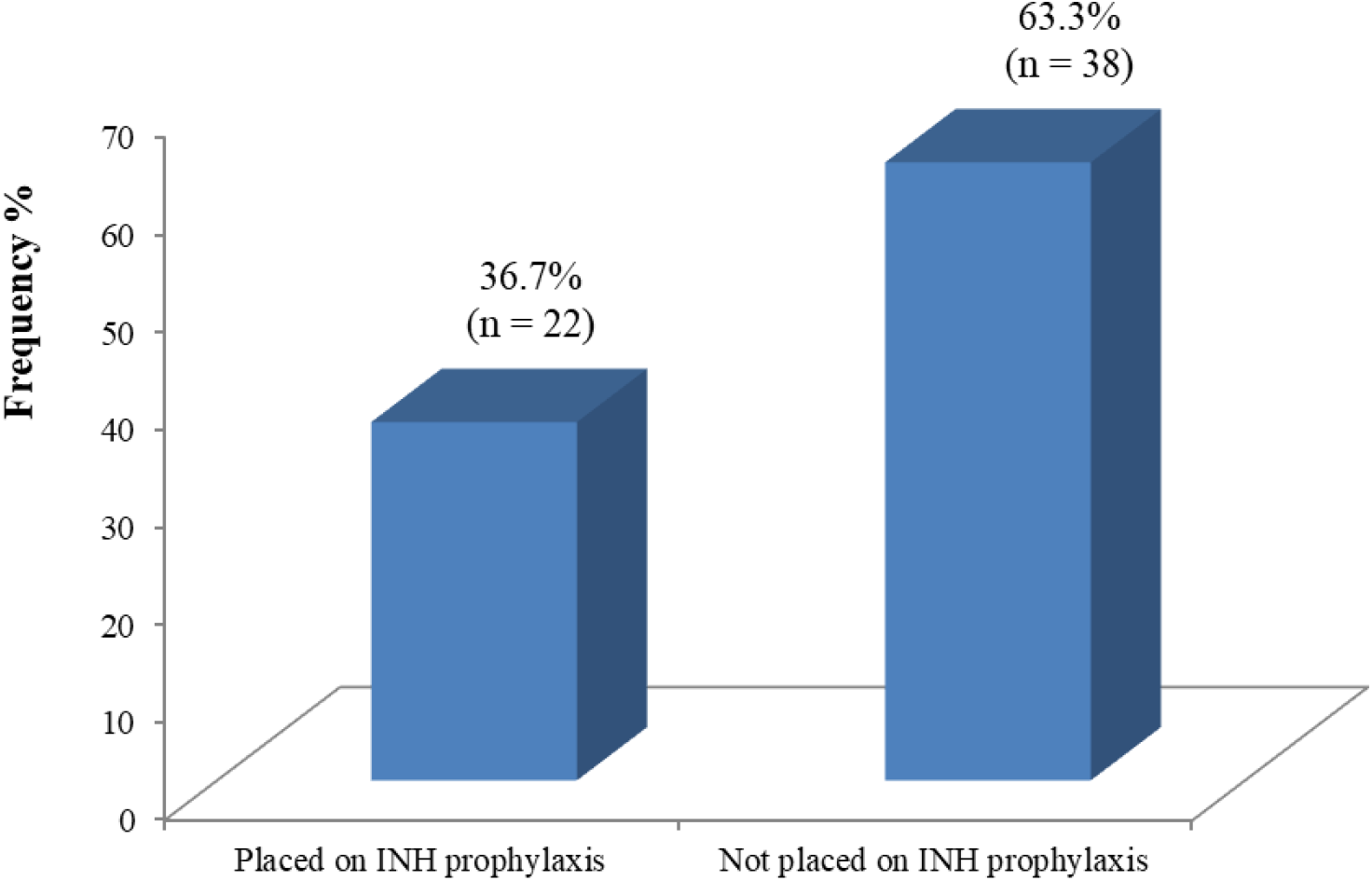
Distribution of PLWH in Obio-Akpor placed on INH prophylaxis

## DISCUSSION

### The proportion of HIV-positive Patients Screened for TB

Guideline for TB preventive therapy in several regions recommends the screening of TB among PLWH during each clinical visit (Fomundam et al., 2020). Utilising ACF has been identified as an effective strategy in enhancing TB control activities (Kempker et al., 2021), while Igbokwe et al. (2020) reported that ACF is very necessary because the risk of progressing from latent to active TB is greater in PLWH than among those without HIV infection. According to the result of this study, the proportion of HIV-positive patients screened for TB was 738 (79.8%) while those who had TB diagnostic evaluation among them was 211 (38.6%). This result is lower than that of Igbokwe et al. (2020) who reported that 98.8% and 80% of PLWH had their TB screening and diagnostic evaluation done and documented at the last visit to the clinic. Pasipamire et al. (2016) also reported that 92.8% of the cases were screened and 54% had their diagnostic evaluation done, while Denegetu and Dolamo (2014), Owiti et al. (2019) and Fomundam et al. (2020) also reported that 97.0%, 83.5% and 94% of PLWH were screened for TB. On the other hand, the result from the study of Galeto et al. (2017) revealed a lower screening rate of 75.2% among PLWH in Harari, Eastern Ethiopia. The differences between the rate of screening for TB among PLWH in the various studies could attributable to the difference in TB case management and the structure of the healthcare management system adopted in the different settings. It could also be linked to possible variations in the study designs, duration, and/or population size applicable to the different studies.

### The proportion of HIV/TB Positive patients placed on INH prophylaxis

The use of IPT has been identified as an effective public health intervention for the prevention of active TB infections among PLWH (Fomundam et al., 2020), while WHO in 2008 recommended the intervention of IPT as a mainstay to reduce incident TB in PLWH in high TB burden countries (WHO, 2008). According to the report of this study, 38.7% of the HIV/TB co-infected patients were placed on INH prophylaxis. Contrary to this finding, Abossie and Yohanes (2017), Igbokwe et al. (2020), Legese et al. (2020) reported that 68.0%, 96.2%, and 62.1% of the PLWH with positive TB evaluation were placed on anti-TB treatment. Other studies in some other SSA regions by Adjobimey et al. (2016) in Benin, Egere et al. (2016) in Gambia, Tadesse et al. (2016) in Ethiopia, and Birungi et al. (2018) in Gambia reported higher rates of IPT initiation (99.0%, 89.0%, 64.3%, and 89.0% respectively) upon testing positive for TB among PLWH. On the other hand, a recent study by Oonyu et al. (2022) in Uganda reported a lower rate of IPT initiation (34.2%). Also, an Indian study by Shivaramakrishna et al. (2014) reported a lower rate of 33.0%, while the studies of Kagujj et al. (2019) and Teklay et al. (2016) reported a lower rate of 30.0% and 20.0% in Zambia and Ethiopia respectively. The differences between the findings in these studies could be linked to the result of the integration of TB programs into the HIV programs in some of the settings or the lack of it, as well as disparities in the study design and sampling techniques.

## Conclusion and Recommendation

The findings from this study have revealed that the proportion of HIV-positive patients screened for TB and had TB diagnostic evaluation in Obio/Akpor LGA of Rivers state, Nigeria was 79.8% and 38.6% respectively, while only 38.7% of the HIV/TB co-infected patients were placed on INH prophylaxis. This shows that insufficient attention is being paid to TB diagnostic evaluation and IPT either due to a lack of appropriate equipment or a lack of sensitization among the healthcare workers (HCWs). Hence, it is recommended that the PHCs in the LGA should be equipped with appropriate devices for TB diagnosis as well as engage the HCWs in sensitization workshops for improved diagnosis and management of TB case, especially among PLWH.

## Data Availability

All data produced in the present study are available upon reasonable request to the authors

